# Automation risk and subjective wellbeing in the UK

**DOI:** 10.1101/2024.01.18.24301484

**Authors:** Jiyuan Zheng, Bertha Rohenkohl, Mauricio Barahona, Jonathan M Clarke

## Abstract

The personal well-being of workers may be influenced by the risk of job automation brought about by technological innovation. Here we use data from the Understanding Society survey in the UK and a fixed-effects model to examine associations between working in a highly automatable job and life and job satisfaction. We find that employees in highly automatable jobs report significantly lower job satisfaction, a result that holds across demographic groups categorised by gender, age and education, with higher negative association among men, higher degree holders and younger workers. On the other hand, life satisfaction of workers is not generally associated with the risk of job automation, a result that persists among groups disaggregated by gender and education, but with age differences, since the life satisfaction of workers aged 30 to 49 is negatively associated with job automation risk. Our analysis also reveals differences in these associations across UK industries and regions.

## 1. Introduction

Work is a major part of the lives of people of working age across the world. It has been estimated that people spend between 30 and 50% of their waking hours at work and the strong relationship between job satisfaction and overall wellbeing is well known (Layard & de Neve, 2023a, Rohenkohl & Clarke 2023).

For those in employment, job satisfaction can be profoundly affected by the characteristics of the workplace. Good interpersonal relationships with colleagues, having interesting work, and job security may lead to higher job satisfaction, while jobs that are difficult, stressful or offer a poor work-life balance may lower job satisfaction (de Neve, 2018). Furthermore, unemployment is consistently associated with lower wellbeing, and the experience of being unemployed can have lasting negative effects on wellbeing even after finding a new job (Layard & de Neve, 2023b). Therefore, changes to labour markets that alter the availability or characteristics of work across industries may in turn have important and widespread consequences the wellbeing of societies as a whole.

The emergence of new automation technologies, including artificial intelligence and robotics in recent years, have the potential to significantly change the world of work (Acemoglu & Restrepo, 2018). These technologies are already being applied across industries and may both substitute and complement existing occupations. Their introduction may lead to the loss of jobs in some occupations and the creation of entirely new occupations, and may also change the tasks an employee performs, such as the automation of more routine tasks, leaving employees to focus on more creative and rewarding tasks (Nazareno & Schiff, 2021).

While extensive research has been conducted on how labour markets respond to new technologies, much of this has focussed on their impact on employment, wages and productivity. These studies indicate a complex array of effects on labour markets in which both increases and decreases in unemployment and wages may occur differently across industries and over time (Acemoglu & Restrepo, 2018, Rohenkohl & Clarke, 2023).

Less is known about the impact of the emergence of automation technologies on the job satisfaction and overall life satisfaction of workers. Recent studies have examined various aspects of the relationship between the risk of job automation and the subjective job satisfaction or life satisfaction of workers across different countries (Johnson et al., 2020; Tamers et al., 2020; Schwabe & Castellacci, 2020; Lordan & Stringer, 2022; Liu, 2022; Gorny & Woodard, 2020; Nazareno & Schiff, 2021; Stankevičiūtė, Staniškienė & Ramanauskaitė, 2021), albeit not focussed on the United Kingdom.

Just as there is no consensus on the impact of automation technologies on labour markets, there is no consensus on how job automation may affect the subjective wellbeing of workers. Research has demonstrated that fear of robot technologies is associated with lower levels of life satisfaction (Hinks, 2021), and fear of replacement by smart machines has a negative impact on job satisfaction (Schwabe & Castellacci, 2020). Existing literature has also suggested that lower job satisfaction is related to higher awareness of automation technologies (Brougham & Haar, 2018). On the other hand, automation may enhance wellbeing by reducing workload, lowering stress and improving work-life balance (Lordan & Stringer, 2022; Makridis & Han, 2021). Indeed, a rise in technological growth has been shown to correlate with increased current and expected future life satisfaction (Makridis & Han, 2021).

In the UK, around 30% of occupations are at high risk of being automated, with jobs in retail, manufacturing, administration and logistics thought to face the highest job automation risk (Berriman & Hawksworth, 2017). Here, we investigate the relationship between job automation risk and the reported job and life satisfaction of UK employees using data collected over successive waves of the Understanding Society (USoc) survey (University of Essex, Institute for Social and Economic Research, 2022) between 2009 and 2022. Life satisfaction and job satisfaction responses from the survey are used as measures of subjective wellbeing, whereas the automation risk is quantified by the probability of computerisation of a worker’s current occupation, as estimated by Frey & Osborne (2017). We then apply a fixed-effects model with individual-, wave- and region-specific fixed effects to establish he presence of significant associations.

We find that employees in highly automatable occupations report significantly lower job satisfaction, a finding confirmed for both men and women, with a stronger negative association among men. Age-wise, this negative association is present in those aged below 50 years of age, but no association is observed for older employees. Additionally, workers with a degree or other higher degree in highly automatable jobs report lower levels of job satisfaction compared with those without a degree or other higher degree. On the other hand, our analysis finds no significant relationship between job automation risk and the overall life satisfaction of employees, a finding reproduced across groups disaggregated by gender and education levels. Age-wise, life satisfaction is negatively associated with the risk of job automation only for people aged 30-49. Finally, our results disaggregated by industries indicate that working in a highly automatable occupation is negatively related to job satisfaction in the agriculture, manufacturing, transport and service industries.

## 2. Data and Methods

### 2.1. Data

This paper uses UK data from the Understanding Society survey (USoc) led by the University of Essex, Institute for Social and Economic Research. The sample is nationally representative of the population in the UK^2^, and respondents are interviewed yearly if they live in the UK and agree to take part (Institute for Social and Economic Research, 2021). The Understanding Society survey contains rich information on a wide range of socio-demographic characteristics, as well as work and wellbeing across the UK (University of Essex, Institute for Social and Economic Research, 2022).

We use data from wave 1 (January 2009-March 2011) to wave 12 (January 2020-May 2022). The sample is made up of individuals aged 16 years and above who are in employment at the time of the survey. Self-employed respondents are excluded due to missing data on job-related characteristics for this group of workers. To measure the automation risk of each occupation, we focus on employees whose three-digit Standard Occupational Classification (SOC) 2000 code can be matched to occupations whose probabilities of job automation have been estimated by Frey and Osborne (2017). We include only respondents with non-missing data for all measures of subjective wellbeing including life satisfaction and job satisfaction resulting in a dataset of 43471 workers across 12 waves.

#### 2.1.1. Subjective Wellbeing

Subjective wellbeing is a synoptic reflection of how individuals feel about their own lives, and thus does not rely on analysts deciding what specific factors should be used to quantify wellbeing (Hicks, Tinkler & Allin, 2013). Here, we use a widely adopted measure of subjective wellbeing: life satisfaction. National life satisfaction statistics have been adopted as a measure of wellbeing of citizens to inform policy in the UK (Diener, Inglehart & Tay, 2013). The life satisfaction variable is created here from the question within USoc about an individual’s overall satisfaction with life, and measured on a seven-point scale, ranging from 1 (“completely dissatisfied”) to 7 (“completely satisfied”). The variable is dichotomized into “not satisfied” (comprising scales 1-4: “completely dissatisfied”, “mostly dissatisfied”, “somewhat dissatisfied”, “neither satisfied nor dissatisfied”) and “satisfied” (comprising scales 5-7: “somewhat satisfied”, “mostly satisfied”, “completely satisfied”).^3^ We then explore whether job automation risk is linked with specific facets of life satisfaction, including satisfaction with health, income and the amount of leisure time, each of which are also included as similar ordinal variables within USoc.

As the second measure of subjective wellbeing, we use job satisfaction. Job satisfaction is typically positively correlated with the subjective wellbeing of those in employment (Bowling, Eschleman & Wang, 2010) and may thus be understood as a component of overall life satisfaction. In USoc, the survey question concerning job satisfaction asks how satisfied the respondent is with the present job on a seven-point scale, which is dichotomized between ‘satisfied’ (1-4) or ‘not satisfied’ (5-7). We adopt the dichotomized variable for life satisfaction and job satisfaction for clarity of exposition but the results hold without change for the seven-point variable (see section 2.2.1).

#### 2.1.2. Automation Risk

As a measure of automation risk, we use the probability of future computerisation for 702 occupations from Frey and Osborne (2017) (hereafter FO). Based on the 2010 version of O*Net data, which includes characteristics of 903 occupations^4^ in the US, FO classify 70 occupations as fully automatable or non-automatable and implement a classification algorithm to derive the probability of computerisation of all occupations based on features such as perception and manipulation, creativity and social intelligence required to carry out job tasks. The FO measure thus indicates the automatability of jobs beyond the computerisation of routine tasks (Frey & Osborne, 2017). To match the FO probabilities of computerisation to the three-digit SOC 2000 occupation codes in USoc, the probability of job automation corresponding to four-digit UK SOC 2010 codes is obtained from ONS (White, Lacey & Ardanaz-Badia, 2019) and crosswalked to four-digit UK SOC 2000 codes. Then, we assign the mean probability of job automation to the corresponding three-digit SOC 2000. To make our results directly comparable to other work (e.g., Adamczyk, Monasterio & Fochezatto (2021), Albuquerque et al. (2019)), we create a binary variable that equals 1 when the probability of automation is in the top quartile (i.e., larger than the 75^th^ percentile of all jobs) and zero otherwise. However, our results do not change if the threshold is varied, or if we use the probability of automation as a continuous variable.

### 2.2. Estimation Strategy

Our data captures not only variation across individuals, but also changes within individuals over time, which can help identify unobserved heterogeneity affecting outcomes of interest (Hsiao, 1985). Hence, we apply the following fixed effects model to estimate the association between wellbeing and job automation risk, which allows us to remove bias due to possible confounders:

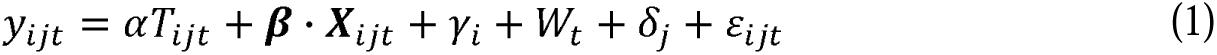

where *y_ijt_* denotes the wellbeing variable (job satisfaction or life satisfaction) of individual *i* in area *j* during wave *t; T_ijt_* is a binary variable which equals one if an employee is working in a highly automatable job and zero otherwise (as described above); *γ_i_* denotes individual fixed effects; *W_t_* is the wave fixed effects; *δ_j_* represents the region fixed effects and *ε_ijt_* is the idiosyncratic error term. The vector of control variables ***X****_ijt_* includes socio-demographic characteristics (i.e., age, gender, marital status^5^, education levels^6^, ethnicity^7^, the presence of children, housing tenure and rural/urban residence) as well as job-related attributes (i.e., industry classifications^8^, size of workplace^9^, contract type and public/private sector).

The coefficient of interest is a, which indicates the relationship between working in a highly automatable job on wellbeing outcomes. The individual fixed effects capture personal attributes which remain unchanged over time. The wave fixed effects represent factors which could vary over time, such as changes in macroeconomic conditions as well as national health and wellbeing policies. The region fixed effects indicate region-specific features, such as regional economic development, local labour market conditions and local public policies about employment. Standard errors were clustered at the individual level. The model thus removes unobserved individual-, time- and region-specific heterogeneity which may otherwise bias our results.

#### 2.2.1 Robustness

We have confirmed the robustness of our results to the definition of the variables. First, we check that the results do not change for different thresholds when dichotomising the job automation risk into a binary variable, and when we use directly the original automation risk probabilities as a continuous variable. Second, the results remain unchanged when considering the original categorical variables of job satisfaction and life satisfaction on the 7-point Likert type scale analysed through a fixed effect ordered logit model (Baetschmann et al., 2020), which considers unobservable fixed effects in the ordered logit model.

## 3. Results

### 3.1. Descriptive Statistics

Figure A-1 in the Appendix shows the distribution of the probability of job automation over the whole sample. Overall, the probability of automation ranges between 0.2 and 0.7, and we classify the top quartile (21 out of 81 occupations) as highly automatable, so that 16217 out of 43471 respondents work in highly automatable occupations. Table 1 presents descriptive statistics of the sample. Statistically significant differences between those working in highly automatable or less automatable jobs were found for all descriptors, with the exception of the presence of children aged 15 years or under in the household, as shown in Table 1. Notable differences between groups include 22.6% of those working in a highly automatable job having a university degree, compared to 52.6% of those working in a less automatable job. Those working in highly automatable jobs were also more likely than those in less automatable jobs to have worked in the private sector (89.9% vs. 68.8%) and less likely to have worked in the service sector (50.8% vs. 66.8%).

**Table 1.**
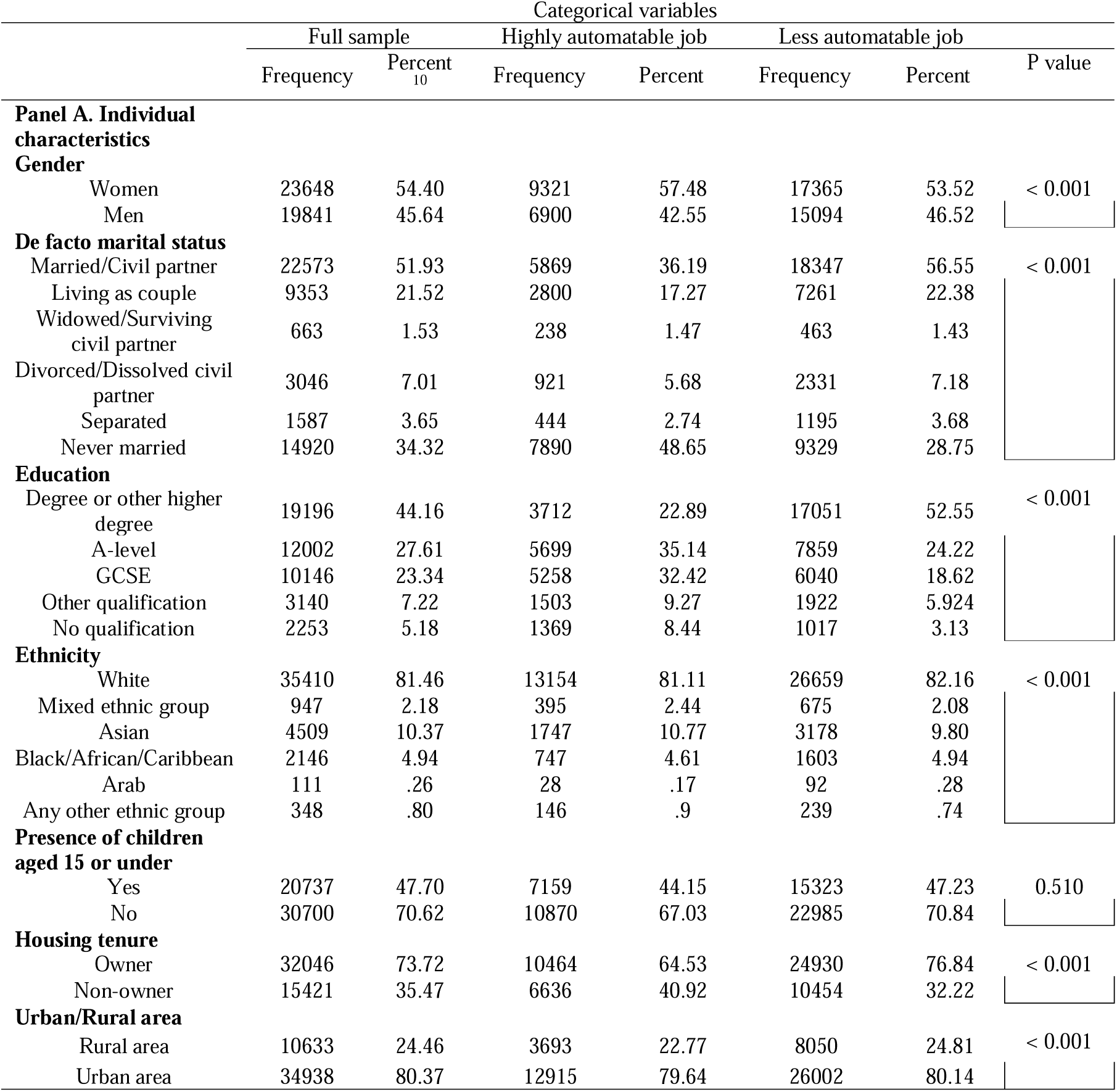
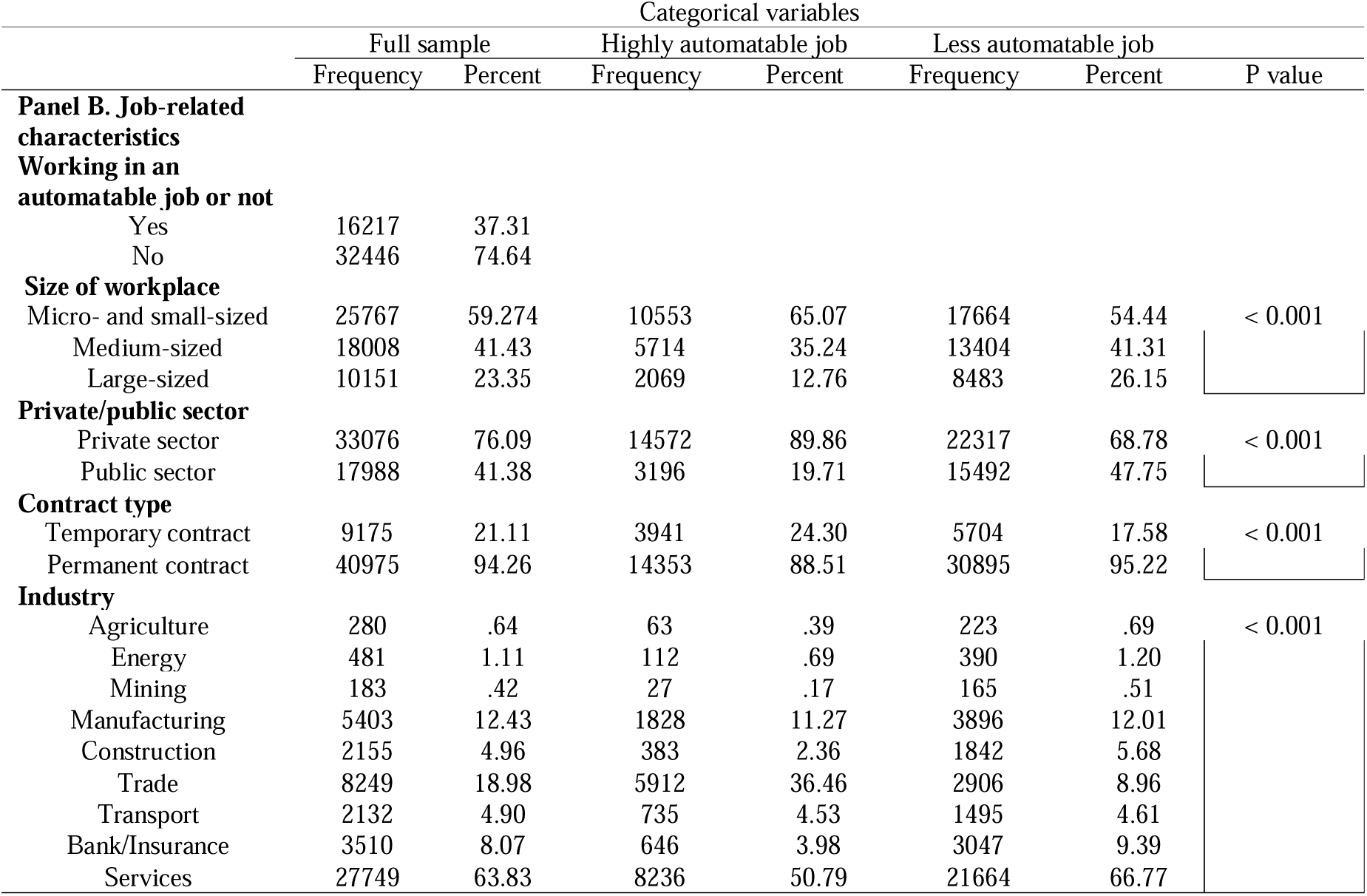
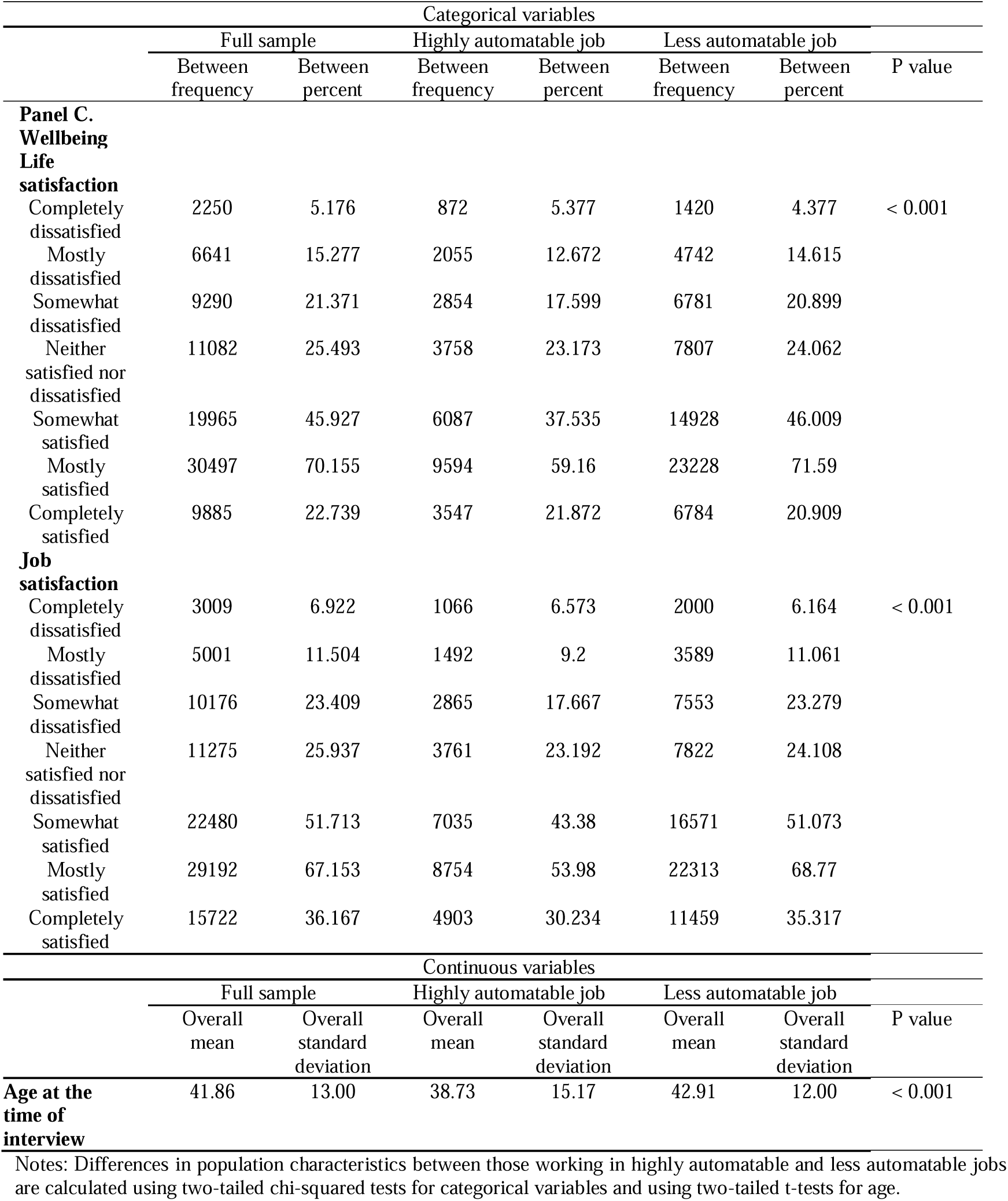
Descriptive statistics. The characteristics of those working in highly automatable and less automatable jobs were compared using chi-squared tests for each of the categorical variables and a t-test for age, the only continuous variable.

Table A-1 in the Appendix compares the distribution of gender, ethnicity, age groups and qualifications between the USoc sample in our empirical study and the population of the UK according to the 2011 UK Census (UK Data Service, 2014). Overall, the distribution of our sample is broadly similar to that of the Census with respect to ethnicity, gender and age groups, yet there is a modest overrepresentation in our sample of individuals with higher qualification, including those holding a degree or other higher degree (level 4+) and GCSE (level 3), while there is an underrepresentation of people with qualifications at level 1 and without qualifications.

### 3.2. Automation Risk and Subjective Wellbeing

Table 2 shows the results of the fixed effects model (1) for the full sample. We find that working in a highly automatable job is associated with a reduction in the level of job satisfaction. On the other hand, we find no evidence of a significant relationship between the life satisfaction of workers and automation risk. Yet we find significant associations with specific dimensions of life satisfaction; as shown in Table 3, working in a job at high risk of automation is associated with a small decline in levels of satisfaction with health, and a larger reduction in the level of satisfaction with income; but an increase in the level of satisfaction with the amount of leisure time.

**Table 2.** The relationship between working in a highly automatable job and life satisfaction/job satisfaction.

|  | (1)<br>Life<br>satisfaction | (2)<br>Job<br>satisfaction |
| --- | --- | --- |
| **Full sample** |  |  |
| Highly automatable job | -0.00420<br>(0.00528) | -0.0490***<br>(0.00625) |
| Mean of Dep. Var. | 0.767 | 0.787 |
| Number of Obs | 187630 | 187630 |
| Number of Individuals | 32468 | 32468 |

Notes: The fixed effect model in Equation 1 is applied, which includes all sociodemographic and job-related control variables as well as individual, region and wave fixed effects. Standard errors in parentheses are clustered at the individual level. \*\*\* p < 0.01, \*\* p < 0.05, \* p < 0.1.

**Table 3.** The relationship between working in a highly automatable job and different aspects of life satisfaction.

|  | Different dimensions of life satisfaction |  |  |
| --- | --- | --- | --- |
|  | (1)<br>Satisfaction with<br>health | (2)<br>Satisfaction with<br>income | (3)<br>Satisfaction with<br>amount of leisure<br>time |
| Highly automatable job | -0.00988*<br>(0.00565) | -0.0269***<br>(0.00611) | 0.0220***<br>(0.00629) |
| Mean of Dep. Var. | 0.682 | 0.616 | 0.552 |
| Number of Obs | 187518 | 187476 | 187557 |
| Number of Individuals | 32454 | 32438 | 32457 |

Notes: The fixed effect model in Equation 1 is applied, which includes all sociodemographic and job-related control variables as well as individual, region and wave fixed effects. Standard errors in parentheses are clustered at the individual level. \*\*\* p < 0.01, \*\* p < 0.05, \* p < 0.1.

### 3.3. Subgroup Analysis

In Table 4, we investigate the heterogeneous relationship between job automation risk and life and job satisfaction, by examining subsamples classified by gender, age and education. The results for the gender and education subgroups are fully consistent with the associations found for the entire sample (see Table 2). However, some differences are found when the sample is divided into four age groups (16-29, 30-49, 50-64 and 65+). In contrast to the full sample, workers aged 30-49 in highly automatable jobs experience lower life satisfaction; and no significant (negative) relationship between job automatability and job satisfaction is observed for those aged 50-64 and 65+. Hence the negative associations between risk of automation and job and life satisfaction are stronger in the younger segments of the population.

**Table 4.** Heterogeneous relationship between working in a highly automatable job and life satisfaction/job satisfaction.

|  | (1)<br>Life satisfaction | (2)<br>Job satisfaction |
| --- | --- | --- |
| **Gender** |  |  |
| **Men** |  |  |
| Highly automatable job | -0.00459<br>(0.00842) | -0.0660***<br>(0.0101) |
| Mean of Dep. Var. | 0.772 | 0.772 |
| Number of Obs | 82845 | 82845 |
| Number of Individuals | 14540 | 14540 |
| **Women** |  |  |
| Highly automatable job | -0.00391<br>(0.00677) | -0.0373***<br>(0.00789) |
| Mean of Dep. Var. | 0.764 | 0.799 |
| Number of Obs | 104772 | 104772 |
| Number of Individuals | 17933 | 17933 |
| **Age Group** |  |  |
| **16-29** |  |  |
| Highly automatable job | -0.00274<br>(0.00866) | -0.0740***<br>(0.0103) |
| Mean of Dep. Var. | 0.782 | 0.780 |
| Number of Obs | 35085 | 35085 |
| Number of Individuals | 9325 | 9325 |
| **30-49** |  |  |
| Highly automatable job | -0.0225**<br>(0.00914) | -0.0475***<br>(0.0109) |
| Mean of Dep. Var. | 0.768 | 0.788 |
| Number of Obs | 90038 | 90038 |
| Number of Individuals | 16880 | 16880 |
| **50-64** |  |  |
| Highly automatable job | 0.00315<br>(0.0134) | -0.0145<br>(0.0146) |
| Mean of Dep. Var. | 0.751 | 0.781 |
| Number of Obs | 53813 | 53813 |
| Number of Individuals | 10569 | 10569 |
| **>=65** |  |  |
| Highly automatable job | 0.0616 | 0.0169 |
|  | (0.0503) | (0.0385) |
| Mean of Dep. Var. | 0.840 | 0.914 |
| Number of Obs | 4417 | 4417 |
| Number of Individuals | 1138 | 1138 |
| **Education level** |  |  |
| People with a degree or other higher degree |  |  |
| Highly automatable job | -0.0132 | -0.0647*** |
|  | (0.00938) | (0.0116) |
| Mean of Dep. Var. | 0.795 | 0.793 |
| Number of Obs | 88548 | 88548 |
| Number of Individuals | 14698 | 14698 |
| People without a degree or other higher degree |  |  |
| Highly automatable job | -0.00391 | -0.0427*** |
|  | (0.00668) | (0.00783) |
| Mean of Dep. Var. | 0.743 | 0.782 |
| Number of Obs | 97929 | 97929 |
| Number of Individuals | 18458 | 18458 |

Notes: The fixed effect model in Equation 1 is applied, which includes all sociodemographic and job-related control variables as well as individual, region and wave fixed effects. Standard errors in parentheses are clustered at the individual level. \*\*\* $p < 0.01$ , \*\* $p < 0.05$ , \* $p < 0.1$ .

To further quantify the differences in associations between the subgroups in Table 4, we added interaction terms between job automation and particular variables for gender, education or age (see Table 5). The most salient differences are as follows. Women exhibit a significantly less negative association between job satisfaction and job automatability, as compared to men. In contrast, workers with a degree or other higher degree report a significantly more negative association between job satisfaction and risk of job automation, as compared to people without a degree. Relative to the age group 30-49, workers aged 16-29 report a significantly more negative association between job satisfaction and risk of job automation, but the opposite applies to older workers (above 50). The negative association between risk of job automation and life satisfaction found in the age group 30-49 is found to be significantly more negative than in any of the other age groups.

**Table 5.** The comparison of the relationship between working in a highly automatable job and life satisfaction and job satisfaction across subgroups.

|  | (1) | (2) |
| --- | --- | --- |
|  | Life satisfaction | Job satisfaction |
| **Comparison between genders:** |  |  |
| Women × Highly automatable job | -0.00241 | 0.0367*** |
|  | (0.0103) | (0.0123) |
| Mean of dep. var. | 0.767 | 0.787 |
| Number of obs | 187630 | 187630 |
| Number of individuals | 32468 | 32468 |
| **Comparison between age groups:** |  |  |
| 16-29 × Highly automatable job | 0.0166* | -0.0442*** |
|  | (0.00928) | (0.0105) |
| 50-64 × Highly automatable job | 0.0235** | 0.0164* |
|  | (0.00963) | (0.00987) |
| >=65 × Highly automatable job | 0.0328 | 0.0230 |
|  | (0.0208) | (0.0206) |
| Mean of dep. var. | 0.767 | 0.787 |
| Number of obs | 187630 | 187630 |
| Number of individuals | 32468 | 32468 |
| **Comparison between levels of qualification:** |  |  |
| Degree or other higher degree × Highly automatable job | -0.0121 | -0.0303** |
|  | (0.0102) | (0.0122) |
| Mean of dep. var. | 0.767 | 0.787 |
| Number of obs | 187630 | 187630 |
| Number of individuals | 32468 | 32468 |

Finally, Table 6 explores variability by industry sectors. The results on life satisfaction are consistent with the full sample, i.e., there is no significant relationship between automation risk and life satisfaction across all industries. On the other hand, while some industries exhibit the same negative association between job automation risk and job satisfaction as in the full sample (i.e., agriculture, manufacturing, transport and services), there is no significant (negative) association for the other sectors (i.e., energy, mining, construction, trade, bank/insurance). For regional effects, see Table A-2 in the Appendix.

**Table 6.** The relationship between working in a highly automatable job and life satisfaction/job satisfaction across different industries in the UK.

|  | Life satisfaction |  |  |  |  |  |  |  |  |
| --- | --- | --- | --- | --- | --- | --- | --- | --- | --- |
|  | (1)<br>Agriculture | (2)<br>Energy | (3)<br>Mining | (4)<br>Manufacturing | (5)<br>Construction | (6)<br>Trade | (7)<br>Transport | (8)<br>Bank/Insurance | (9)<br>Services |
| Highly automatable job | -0.124<br>(0.132) | -0.0686<br>(0.0595) | -0.00477<br>(0.0638) | -0.0319<br>(0.0208) | 0.0590<br>(0.0446) | -0.00914<br>(0.0160) | -0.0490<br>(0.0361) | -0.0314<br>(0.0270) | -0.00176<br>(0.00810) |
| Mean of Dep. Var. | 0.792 | 0.800 | 0.837 | 0.775 | 0.789 | 0.734 | 0.739 | 0.795 | 0.771 |
| Number of Obs | 717 | 1686 | 645 | 19659 | 6581 | 23248 | 6793 | 12279 | 111530 |
| Number of Individuals | 161 | 333 | 132 | 3732 | 1385 | 5078 | 1364 | 2362 | 20100 |
|  | Job satisfaction |  |  |  |  |  |  |  |  |
|  | (1)<br>Agriculture | (2)<br>Energy | (3)<br>Mining | (4)<br>Manufacturing | (5)<br>Construction | (6)<br>Trade | (7)<br>Transport | (8)<br>Bank/Insurance | (9)<br>Services |
| Highly automatable job | -0.345*<br>(0.188) | -0.0766<br>(0.0905) | 0.0745<br>(0.0796) | -0.0745***<br>(0.0226) | -0.0627<br>(0.0583) | -0.0229<br>(0.0175) | -0.130***<br>(0.0380) | -0.0359<br>(0.0310) | -0.0406***<br>(0.00940) |
| Mean of Dep. Var. | 0.900 | 0.795 | 0.816 | 0.776 | 0.797 | 0.762 | 0.771 | 0.785 | 0.797 |
| Number of Obs | 717 | 1686 | 645 | 19659 | 6581 | 23248 | 6793 | 12279 | 111530 |
| Number of Individuals | 161 | 333 | 132 | 3732 | 1385 | 5078 | 1364 | 2362 | 20100 |

Notes: The fixed effect model in Equation 1 is applied, which includes all sociodemographic and job-related control variables as well as individual, region and wave fixed effects. \*\*\* p < 0.01,
\*\* p < 0.05, \* p < 0.1.

### 3.4. Robustness Checks

We have conducted additional analyses to confirm the robustness of our results. Firstly, our findings are robust to different definitions of jobs at high risk of automation. Table A-3 shows that the results are consistent when we use the first or second quartile as cut-offs to dichotomise the variable into high- and low-risk, instead of the third quartile. Furthermore, Table A-4 confirms the results if we use the probability of automation as a continuous variable. Secondly, the results are also consistent if we use a seven-point Likert-type scale to define job and life satisfaction and apply a fixed-effects ordered logit model (Baetschmann et al., 2020), as shown in Table A-5.

## 4. Discussion

Using data from the Understanding Society survey from 2009-22, we find a significant association between working in a job at high risk of automation and lower job satisfaction in UK workers, but no consistent relationship to their overall life satisfaction. These findings agree with results for the USA (Nazareno & Schiff, 2021; Liu, 2022), Australia (Lordan & Stringer, 2022) and 29 European countries (Gorny & Woodard, 2020).

The negative relationship we find between job automation risk and job satisfaction may be interpreted as the result of job insecurity induced by the pre-existing or impending introduction of automation technologies perceived by workers to pose a risk to their jobs. Echoing findings from Nazareno & Schiff, 2021 and Lordan & Stringer, 2022, job insecurity may lead to lower job satisfaction through increased stress at work and uncertainty about future employment. Job insecurity is an established cause of lower work-related and general wellbeing, which if chronic may in turn lead to persistent lower wellbeing and a deterioration in mental and physical health. (de Witte, Pienaar & de Cuyper, 2016).

On the other hand, the lack of a significant association between working in a highly automatable job and life satisfaction can be qualified by the observed differences across the three dimensions of life satisfaction (Table 3), where we find a significant negative association with satisfaction with income (a job-related dimension) compensated by a significant positive association with satisfaction with the amount of leisure time. Where this is the result of existing implementation of automation technologies, this finding may indicate a transition to shorter working hours for lower pay. Conversely, where this is in anticipation of the introduction of automation technologies, these findings may indicate a greater emphasis on time spent outside of work driven by employees ‘quietly quitting’ in the face of job insecurity.

Our analysis of demographic subgroups also reveals differential trends. The negative relationship of risk of job automation with both life satisfaction and job satisfaction is only observed among age groups 16-29 and, especially, 30-49. This broadly aligns with Schwabe & Castellacci (2020), who suggest that younger employees may consider job replacement by automation technologies a potential threat to future employment prospects while older workers may expect these changes to occur beyond their working lifetimes. Additionally, younger workers are more likely to be aware of the potentially negative consequences of the adoption of new technologies for their future employment (Brougham & Haar, 2018). The relatively greater magnitude of the negative relationship between automation risk and job satisfaction among employees with a degree or other higher degree could reflect higher expectations for their jobs given the greater investment of time and energy in their human capital (Schwabe & Castellacci, 2020).

We also find evidence of lower job satisfaction among employees working in highly automatable jobs in agriculture, manufacturing, transport and services. Occupations in these industries are particularly susceptible to automation according to Frey & Osborne (2017), which may lead to greater collective awareness among workers of the potential for automation technologies to transform their work and leading to lower job satisfaction primarily for the reasons of job insecurity described above (Brougham & Haar, 2018).

There are several limitations in our study. Firstly, we use automation risk, as defined by Frey & Osborne (2017), to understand the extent to which automation technologies may have influenced the working lives of respondents. Despite being a widely used reference measure of automation risk, FO cannot differentiate employees working in roles already influenced by automation technologies from those where such technologies have not been implemented. As such, our measure of occupational automation risk unavoidably captures a mixed picture of existing, anticipated and unanticipated exposure to automation technologies by workers.

The FO measures, in being limited to occupations that can be automated by computer-controlled equipment, capture a narrower range of occupations than may be affected by automation technologies. Furthermore, the six years since the publication of the FO measures, and the ten years since the original primary data collection (Frey and Osborne, 2013), represent a long time when considering technological change. Our sample also excludes self-employed individuals due to the unavailability of job-related data, an area that deserves further research. Finally, our use of retrospective survey data and estimates of occupational automation risk by necessity exclude more recent technological advances including the advent of large language models such as Chat GPT, for which no data are yet available.

Collectively, our findings indicate a complex, multifactorial relationship between occupational automation risk, job satisfaction and wellbeing. Whether through anticipated or existing exposure to automation technologies at work, it is younger workers and those working in the most automatable industries who have the most negative associations between occupational automation risk and job satisfaction. More broadly, life satisfaction appears less sensitive to occupational automation risk, pointing to varying perceptions of the importance of work in life. Further empirical research is required to determine the specific mechanisms underlying the negative relationship between job automation and subjective wellbeing. These could include higher levels of job insecurity, higher cognitive demands arising from the automation of less cognitively demanding tasks, loss of meaningfulness of work, or increased surveillance and control in the workplace.

Irrespective of the cause, our analysis identifies existing differences according to the risk of job automation across industries and workers in how satisfying they find their work and to a lesser extent in how this goes on to influence their wider satisfaction with their lives. Directives and institutional policies introduced to ensure a fair transition to the widespread use of automation technologies at work must thus consider differences between individuals and industries, and whether the actual degree of job automation can influence wellbeing outcomes among employees.

## Contributorship statement

JZ: Conceptualisation, Formal Analysis, Writing - Original Draft

BR: Conceptualisation, Formal Analysis, Writing - Review & Editing

MB: Conceptualisation, Writing - Review & Editing, Supervision

JC: Conceptualisation, Writing - Review & Editing, Supervision

## Financial disclosures

None.

## Declaration of competing interest

The authors declare no competing interests in relation to the work.

## Data availability

Understanding Society data used in this study were obtained from the UK Data Service https://beta.ukdataservice.ac.uk/datacatalogue/series/series?id=2000053 Occupational automation risk scores were obtained from the Office for National Statistics https://www.ons.gov.uk/employmentandlabourmarket/peopleinwork/employmentandemployeetypes/datasets/probabilityofautomationinengland

## Acknowledgements

This paper is part of the Pissarides Review of the Future of Work and Wellbeing, based at the Institute for the Future of Work and funded by the Nuffield Foundation. Bertha Rohenkohl, Jiyuan Zheng and Mauricio Barahona acknowledge support from the Nuffield Foundation. Mauricio Barahona also acknowledges support by the EPSRC under grant EP/N014529/1 funding the EPSRC Centre for Mathematics of Precision Healthcare at Imperial. Jonathan Clarke acknowledges support from the Wellcome Trust (215938/Z/19/Z). We would like to thank Professor Christopher Pissarides for his helpful comments on earlier versions of this manuscript.

### Appendix A Tables and Figures

**Figure A-1.**
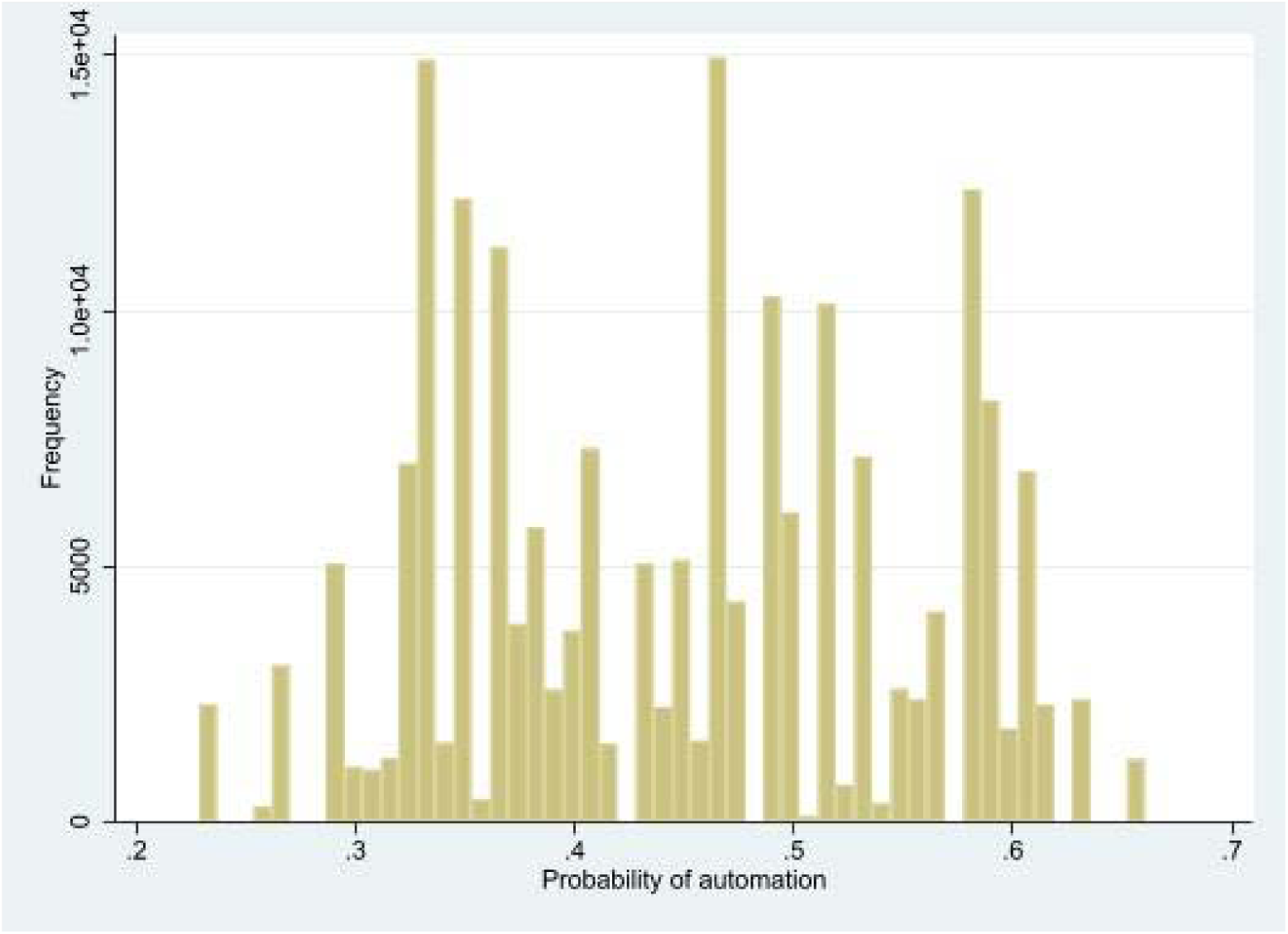
The histogram of the probability of job automation for the entire sample. Notes: The figure displays the histogram of the probability of job automation using measures of job automation risk from Frey & Osborne (2017).

**Table A-1.** Sample representativeness.

|  | Percent (%) from Census | Percent (%) from the sample |
| --- | --- | --- |
| **Ethnicity** |  |  |
| White | 87.166 | 81.46 |
| Asian | 6.922 | 10.37 |
| Black/African/Caribbean | 3.015 | 4.94 |
| Mixed ethnic group | 1.979 | 2.18 |
| Arab and any other ethnic group | 0.919 | 1.06 |
| **Gender** |  |  |
| Women | 50.891 | 54.40 |
| Men | 49.109 | 45.64 |
| **Age groups** |  |  |
| 16-29 | 23.048 | 19.99 |
| 30-49 | 34.281 | 33.54 |
| 50-64 | 22.446 | 24.57 |
| >=65 | 20.225 | 21.89 |
| **NVQ Qualification** |  |  |
| Level 1 | 15.374 | 7.22 |
| Level 2 | 16.558 | 23.34 |
| Level 3 | 13.236 | 27.61 |
| Level 4+ | 29.510 | 44.16 |
| No qualifications | 25.322 | 5.18 |

Notes: This table shows the percentage of population by ethnicity, gender, age group and the highest level of qualifications across the UK based on UK 2011 Census data compared with the sample used in this study.

**Table A-2.**
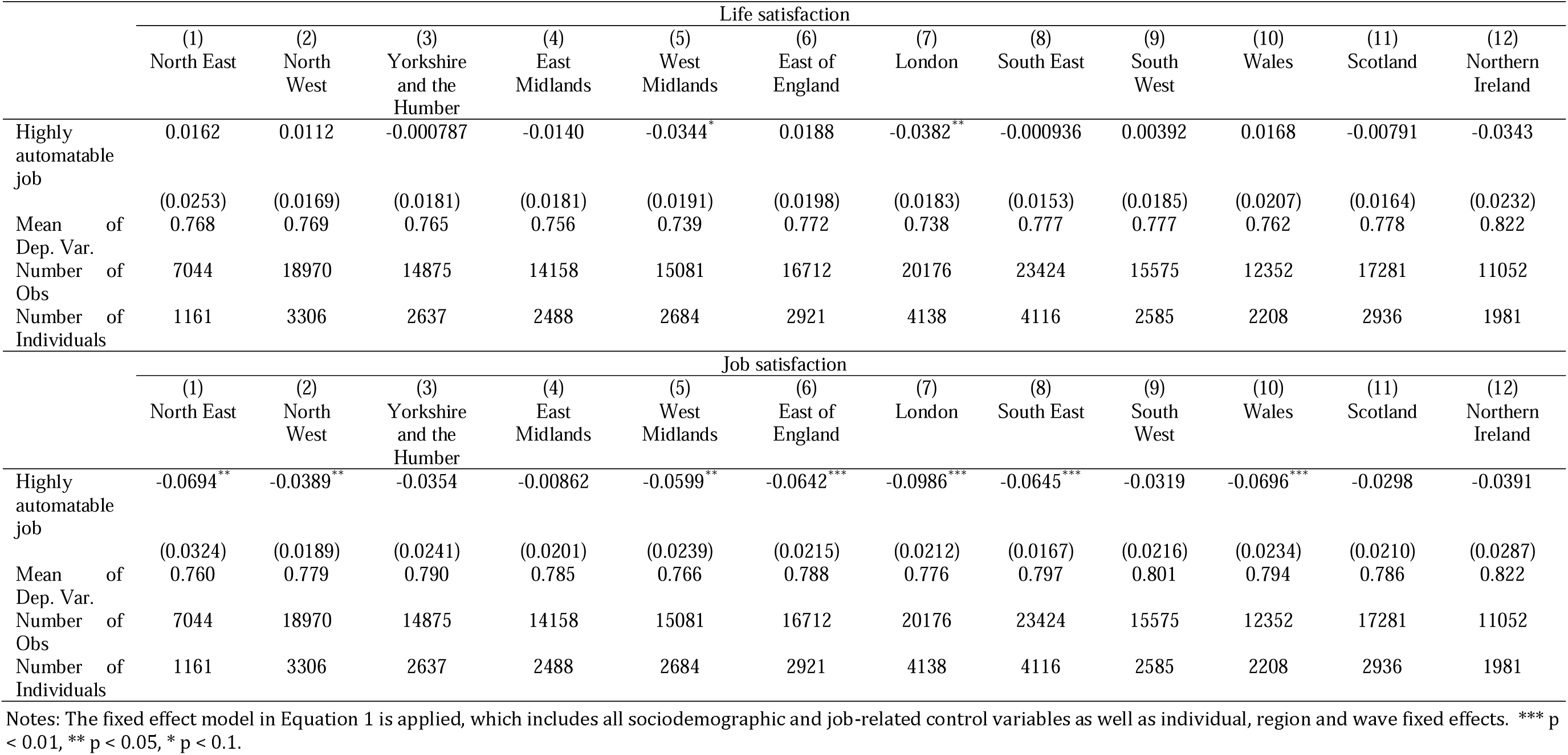
The relationship between working in a highly automatable job and life satisfaction and job satisfaction across regions in the UK.

### Robustness Checks

**Table A-3.** The relationship between working in a highly automatable job with different cut-offs and life satisfaction/job satisfaction.

|  | (1) | (2) |
| --- | --- | --- |
|  | Life satisfaction | Job satisfaction |
| **Probability of automation ≥ 1st quartile** |  |  |
| Working in a highly automatable job | -0.00527<br>(0.00476) | -0.0252***<br>(0.00543) |
| Mean of dep. var. | 0.767 | 0.787 |
| Number of obs | 187630 | 187630 |
| Number of individuals | 32468 | 32468 |
| **Probability of automation ≥ 2nd quartile** |  |  |
| Working in a highly automatable job | -0.00453<br>(0.00463) | -0.0364***<br>(0.00543) |
| Mean of dep. var. | 0.767 | 0.787 |
| Number of obs | 187630 | 187630 |
| Number of individuals | 32468 | 32468 |

\*\*\* $p < 0.01$ .

**Table A-4.** The relationship between the probability of job automation and life satisfaction/job satisfaction.

|  | (1) | (2) |
| --- | --- | --- |
|  | Life satisfaction | Job satisfaction |
| Probability of job automation | -0.0352<br>(0.0244) | -0.266***<br>(0.0285) |
| Mean of Dep. Var. | 0.767 | 0.787 |
| Number of Obs | 187630 | 187630 |
| Number of Individuals | 32468 | 32468 |

\*\*\* $p < 0.01$ .

**Table A-5.** The marginal effect of working in a highly automatable job on life satisfaction and job satisfaction based on the fixed effects ordered logit model.

|  | (1) | (2) |
| --- | --- | --- |
|  | Life satisfaction | Job satisfaction |
| Working in a highly automatable job |  |  |
| Completely dissatisfied | 0.0000534<br>(0.000428) | 0.00478***<br>(0.000673) |
| Mostly dissatisfied | 0.000173<br>(0.00138) | 0.00810***<br>(0.00114) |
| Somewhat dissatisfied | 0.000249<br>(0.00200) | 0.0171***<br>(0.00240) |
| Neither satisfied nor dissatisfied | 0.000234<br>(0.00188) | 0.0150***<br>(0.00211) |
| Somewhat satisfied | 0.000262<br>(0.00210) | 0.0206***<br>(0.00291) |
| Mostly satisfied | -0.000675<br>(0.00542) | -0.0333***<br>(0.00469) |
| Completely satisfied | -0.000296<br>(0.00238) | -0.0322***<br>(0.00454) |
| Number of Obs | 185475 | 183556 |
| Number of Individuals | 31748 | 31262 |

Notes: The table shows estimates of the marginal effect of working in a highly automatable job on life satisfaction and job satisfaction using the fixed effects ordered logit model. \*\*\* p<=0.01.

## Footnotes

2 The representativeness of the sample used in empirical analysis is checked in Section 3.1.

3 Using the original categorical variables on the 7-point Likert type scale as outcome variables, the results are similar to those obtained using dichotomous (‘not satisfied’/’satisfied’) job satisfaction as outcome variables. The same applies to the dichotomous job satisfaction variable below.

4 Jobs under the category of “all other” which do not correspond to the Labour Department’s six-digit Standard Occupational Classification are excluded by Frey and Osborne (2017).

5 The marital status is categorised into married/civil partnership/cohabitation, widowed/divorced/separated and single.

6 We classify the highest educational and vocational qualification into the following categories: degree and other higher degree, A-level or equivalent qualifications, GCSE or equivalent qualifications, other qualification and no qualifications.

7 Based on Government Statistical Service (GSS) harmonised standards on ethnic group from the Office for National Statistics (n.d.), ethnicity is classified into white, mixed ethnicity, Asian/Asian British, black/African/Caribbean/black British, Arab and other ethnic groups.

8 The industry of firms classified using two-digit Cross-National Equivalent File (CNEF) industrial classification is aggregated to one-digit CNEF industrial classification.

9 Based on Organisation for Economic Co-operation and Development (2022), organisations are classified into micro and small organisations (1-49 employees), medium-sized organisations (50-499 employees) and large organisations (>=500 employees).

10 The percent and frequency refer to between percent and between frequency. The between percent denotes the percent of individuals who have ever belonged to a certain category of the variable.

## Notes

### Competing Interest Statement

The authors have declared no competing interest.

### Author Declarations

The study used ONLY openly available human data that were originally located at the UK Data Service (https://beta.ukdataservice.ac.uk/datacatalogue/series/series?id=2000053) and the Office for National Statistics (https://www.ons.gov.uk/employmentandlabourmarket/peopleinwork/employmentandemployeetypes/datasets/probabilityofautomationinengland)

## References

Acemoglu, D. & Restrepo, P. (2018) The Race between Man and Machine: Implications of Technology for Growth, Factor Shares, and Employment. American Economic Review. 108 (6), 1488–1542. doi:10.1257/aer.20160696.

Adamczyk, W.B., Monasterio, L. & Fochezatto, A. (2021) Automation in the future of public sector employment: the case of Brazilian Federal Government. Technology in Society. 67, 101722. doi:10.1016/j.techsoc.2021.101722.

Albuquerque, P.H.M., Saavedra, C.A.P.B., de Morais, R.L. & Peng, Y. (2019) The Robot from Ipanema goes Working: Estimating the Probability of Jobs Automation in Brazil. Latin American Business Review. 20 (3), 227–248. doi:10.1080/10978526.2019.1633238.

Autor, D.H. & Dorn, D. (2013) The Growth of Low-Skill Service Jobs and the Polarization of the US Labor Market. American Economic Review. 103 (5), 1553–1597. doi:10.1257/aer.103.5.1553.

Autor, D.H., Katz, L.F. & Kearney, M.S. (2006) The Polarization of the U.S. Labor Market. American Economic Review. 96 (2), 189–194. doi:10.1257/000282806777212620.

Baetschmann, G., Ballantyne, A., Staub, K.E. & Winkelmann, R. (2020) feologit: A new command for fitting fixed-effects ordered logit models. The Stata Journal. 20 (2), 253–275. doi:10.1177/1536867X20930984.

Berriman, R. & Hawksworth, J. (2017) Will Robots Steal Our Jobs? The Potential Impact of Automation on the UK and Other Major Economies.

Bowling, N.A., Eschleman, K.J. & Wang, Q. (2010) A meta-analytic examination of the relationship between job satisfaction and subjective well-being. Journal of Occupational and Organizational Psychology. 83 (4), 915–934. doi:10.1348/096317909X478557.

Brougham, D. & Haar, J. (2018) Smart Technology, Artificial Intelligence, Robotics, and Algorithms (STARA): Employees’ perceptions of our future workplace. Journal of Management & Organization. 24 (2), 239–257. doi:10.1017/jmo.2016.55.

Clément S. Bellet, Jan-Emmanuel De Neve, George Ward (2023) Does Employee Happiness Have an Impact on Productivity?. Management Science 0(0).

Cobb-Clark, D.A. (2015) Locus of control and the labor market. IZA Journal of Labor Economics. 4 (1), 3. doi:10.1186/s40172-014-0017-x.

Cubel, M., NuevoDChiquero, A., SanchezDPages, S. & VidalDFernandez, M. (2016) Do Personality Traits Affect Productivity? Evidence from the Laboratory. The Economic Journal. 126 (592), 654–681. doi:10.1111/ecoj.12373.

De Neve, J. E. (2018). Work and well-being: A global perspective. Global Happiness and Well-Being Policy Report 2018.

De Witte, H., Pienaar, J. & De Cuyper, N. (2016) Review of 30 Years of Longitudinal Studies on the Association Between Job Insecurity and Health and WellDBeing: Is There Causal Evidence? Australian Psychologist. 51 (1), 18–31. doi:10.1111/ap.12176.

Diener, E., Inglehart, R. & Tay, L. (2013) Theory and Validity of Life Satisfaction Scales. Social Indicators Research. 112 (3), 497–527. doi:10.1007/s11205-012-0076-y.

Frey, C.B. & Osborne, M.A. (2017) The future of employment: How susceptible are jobs to computerisation? Technological Forecasting and Social Change. 114, 254–280. doi:10.1016/j.techfore.2016.08.019.

Georgellis, Y., Tsitsianis, N. & Yin, Y.P. (2009) Personal Values as Mitigating Factors in the Link Between Income and Life Satisfaction: Evidence from the European Social Survey. Social Indicators Research. 91 (3), 329–344. doi:10.1007/s11205-008-9344-2.

Ghislieri, C., Molino, M. & Cortese, C.G. (2018) Work and Organizational Psychology Looks at the Fourth Industrial Revolution: How to Support Workers and Organizations? Frontiers in Psychology. 9. https://www.frontiersin.org/articles/10.3389/fpsyg.2018.02365.

Gorny, P.M. & Woodard, R.C. (2020) Don’t Fear the Robots: Automatability and Job Satisfaction. MPRA Paper. https://ideas.repec.org/p/pra/mprapa/103424.html.

Hicks, S., Tinkler, L. & Allin, P. (2013) Measuring Subjective Well-Being and its Potential Role in Policy: Perspectives from the UK Office for National Statistics. Social Indicators Research. 114 (1), 73–86. doi:10.1007/s11205-013-0384-x.

Highhouse, S. & Becker, A.S. (1993) Facet measures and global job satisfaction. Journal of Business and Psychology. 8 (1), 117–127. doi:10.1007/BF02230397.

Hill, T.D., Davis, A.P., Roos, J.M. & French, M.T. (2020) Limitations of Fixed-Effects Models for Panel Data. Sociological Perspectives. 63 (3), 357–369. doi:10.1177/0731121419863785.

Hinks, T. (2021) Fear of Robots and Life Satisfaction. International Journal of Social Robotics. 13 (2), 327–340. doi:10.1007/s12369-020-00640-1.

Hsiao, C. (1985) Benefits and limitations of panel data. Econometric Reviews. 4 (1), 121–174. doi:10.1080/07474938508800078.

Institute for Social and Economic Research (2021) Understanding Society: Waves 1-11, 2009-2020 and Harmonised BHPS: Waves 1-18, 1991-2009, User Guide.

Johnson, A., Dey, S., Nguyen, H., Groth, M., Joyce, S., Tan, L., Glozier, N. & Harvey, S.B. (2020) A review and agenda for examining how technology-driven changes at work will impact workplace mental health and employee well-being. Australian Journal of Management. 45 (3), 402–424. doi:10.1177/0312896220922292.

Khubchandani, J. & Price, J.H. (2017) Association of Job Insecurity with Health Risk Factors and Poorer Health in American Workers. Journal of Community Health. 42 (2), 242–251. doi:10.1007/s10900-016-0249-8.

Layard R, De Neve J-E. (2023a) The Quality of Work. In: Wellbeing: Science and Policy. Cambridge University Press; 2023:178–201.

Layard R, De Neve J-E. (2023b) Unemployment. In: Wellbeing: Science and Policy. Cambridge University Press; 2023:166–177.

Liu, L. (2022) Job quality and automation: Do more automatable occupations have less job satisfaction and health? Journal of Industrial Relations. 00221856221129639. doi:10.1177/00221856221129639.

Lordan, G. & Stringer, E.-J. (2022) People versus Machines: The Impact of Being in an Automatable Job on Australian Worker’s Mental Health and Life Satisfaction. 101.

Makridis, C.A. & Han, J.H. (2021) Future of work and employee empowerment and satisfaction: Evidence from a decade of technological change. Technological Forecasting and Social Change. 173 (C). https://ideas.repec.org/a/eee/tefoso/v173y2021ics0040162521005953.html.

Nazareno, L. & Schiff, D.S. (2021) The impact of automation and artificial intelligence on worker well-being. Technology in Society. 67, 101679. doi:10.1016/j.techsoc.2021.101679.

Nieß, C. & Zacher, H. (2015) Openness to Experience as a Predictor and Outcome of Upward Job Changes into Managerial and Professional Positions. PLOS ONE. 10 (6), e0131115. doi:10.1371/journal.pone.0131115.

Office for National Statistics (n.d.) Ethnic group, national identity and religion - Office for National Statistics. https://www.ons.gov.uk/methodology/classificationsandstandards/measuringequality/ethnicgr oupnationalidentityandreligion [Accessed: 11 December 2022].

Organisation for Economic Co-operation and Development (2022) Entrepreneurship - Enterprises by business size - OECD Data. http://data.oecd.org/entrepreneur/enterprises-by-business-size.htm [Accessed: 1 December 2022].

Oswald, A.J., Proto, E. & Sgroi, D. (2015) Happiness and productivity. Journal of labor economics. doi:10.1086/681096.

Pehkonen, J., Viinikainen, J., Böckerman, P., Pitkänen, N., Lehtimäki, T. & Raitakari, O. (2019) Health endowment and later-life outcomes in the labour market: Evidence using genetic risk scores and reduced-form models. SSM - Population Health. 7, 100379. doi:10.1016/j.ssmph.2019.100379.

Rohenkohl B. & Clarke J. (2023) What do we know about automation at work and workers’ wellbeing? *Literature Review Working Paper*, Institute for the Future of Work. doi:10.5281/zenodo.8074436.

Schwabe, H. & Castellacci, F. (2020) Automation, workers’ skills and job satisfaction. PLOS ONE. 15 (11), e0242929. doi:10.1371/journal.pone.0242929.

Stankevičiūtė, Ž., Staniškienė, E. & Ramanauskaitė, J. (2021) The Impact of Job Insecurity on Employee Happiness at Work: A Case of Robotised Production Line Operators in Furniture Industry in Lithuania. Sustainability. 13 (3), 1563. doi:10.3390/su13031563.

Tamers, S.L., Streit, J., Pana-Cryan, R., Ray, T., Syron, L., Flynn, M.A., Castillo, D., Roth, G., Geraci, C., Guerin, R., Schulte, P., Henn, S., Chang, C.-C., Felknor, S. & Howard, J. (2020) Envisioning the future of work to safeguard the safety, health, and well-being of the workforce: A perspective from the CDC’s National Institute for Occupational Safety and Health. American Journal of Industrial Medicine. 63 (12), 1065–1084. doi:10.1002/ajim.23183.

UK Data Service (2014) InFuse. 29 May 2014. https://infuse.ukdataservice.ac.uk/ [Accessed: 22 June 2023].

University of Essex, Institute for Social and Economic Research (2022) Understanding Society: Waves 1-12, 2009-2021 and Harmonised BHPS: Waves 1-18, *1991*-2009. 10.5255/UKDA-SN-6614-18.

Viinikainen, J., Böckerman, P., Hakulinen, C., Kari, J.T., Lehtimäki, T., Raitakari, O.T. & Pehkonen, J. (2022) Schizophrenia polygenic risk score and long-term success in the labour market: A cohort study. Journal of Psychiatric Research. 151, 638–641. doi:10.1016/j.jpsychires.2022.05.041.

Wang, H., Cheng, Z., Zhe Wang, B. & Chen, Y. (2021) Childhood left-behind experience and labour market outcomes in China. Journal of Business Research. 132, 196–207. doi:10.1016/j.jbusres.2021.04.014.

White, S., Lacey, A. & Ardanaz-Badia, A. (2019) The probability of automation in England: 2011 and 2017. 25 March 2019. https://www.ons.gov.uk/employmentandlabourmarket/peopleinwork/employmentandemploye etypes/articles/theprobabilityofautomationinengland/2011and2017 [Accessed: 1 March 2023].

